# Building a COVID-19 Vulnerability Index

**DOI:** 10.1101/2020.03.16.20036723

**Authors:** Dave DeCapprio, Joseph Gartner, Carol J. McCall, Thadeus Burgess, Sarthak Kothari, Shaayaan Sayed

## Abstract

COVID-19 is an acute respiratory disease that has been classified as a pandemic by the World Health Organization. Characterization of this disease is still in its early stages; however, it is known to have high mortality rates, particularly among individuals with preexisting medical conditions. Creating models to identify individuals who are at the greatest risk for severe complications due to COVID-19 will be useful for outreach campaigns to help mitigate the disease’s worst effects. While information specific to COVID-19 is limited, a model using complications due to other upper respiratory infections can be used as a proxy to help identify those individuals who are at the greatest risk. We present the results for three models predicting such complications, with each model increasing predictive effectiveness at the expense of ease of implementation.

## I. Introduction

### i. COVID-19 Virus

Coronaviruses (CoV) are a large family of viruses that cause illnesses ranging from the common cold to more severe diseases such as Middle East respiratory syndrome (MERS-CoV) and severe acute respiratory syndrome (SARS-CoV). CoV are zoonotic, meaning they are transmitted between animals and people. Coronavirus disease 2019 (COVID-19) is caused by a new strain discovered in 2019, severe acute respiratory syndrome coronavirus 2 (SARS-CoV-2), that has not been previously identified in humans[1].

COVID-19 is a respiratory infection with common signs that include respiratory symptoms, fever, cough, shortness of breath, and breathing difficulties. In more severe cases, infection can cause pneumonia, severe acute respiratory syndrome, kidney failure, and death.

### ii. Flattening the Curve

On March 11, 2020, the World Health Organization (WHO) declared COVID-19 to be a pandemic[2]. In their press conference, they were clear that pandemic was not a word they used lightly or carelessly or to cause unreasonable fear. They were also clear to highlight that this is the first pandemic to ever be caused by a coronavirus and that all countries can still act to change its course.

Public health and healthcare experts agree that mitigation is required in order to slow the spread of COVID-19 and prevent the collapse of healthcare systems. On any given day, health systems in the United States run close to capacity[3], and so every transmission that can be avoided and every case that can be prevented has enormous impact.

### iii. Identifying Vulnerable People

The risk of severe complications from COVID-19 is higher for certain vulnerable populations, particularly people who are elderly, frail, or have multiple chronic conditions. The risk of death has been difficult to calculate[4], but a small study[5] of people who contracted COVID-19 in Wuhan suggests that the risk of death increases with age, and is also higher for those who have diabetes, heart disease, blood clotting problems, or have shown signs of sepsis. With an average death rate of 1%, the death rate rose to 6% for people with cancer, high blood pressure, or chronic respiratory disease, 7% for people with diabetes, and 10% for people with heart disease. There was also a steep age gradient; the death rate among people age 80 and over was 15% [6].

Identifying who is most vulnerable is not necessarily straightforward. More than 55% of Medicare beneficiaries meet at least one of the risk criteria listed by the US Centers for Disease Control and Prevention (CDC)[7]. People with the same chronic condition don’t have the same risk, and simple rules can fail to capture complex factors like frailty[9] which makes people more vulnerable to severe infections.

## II. Methods

### i. Datasets

Since real-world data on COVID-19 cases are not readily available, the CV19 Index was developed using close proxy events. A person’s CV19 Index is measured in terms of their near-term risk of severe complications from respiratory infections (e.g. pneumonia, influenza). Specifically, 4 categories of diagnoses were chosen from the Clinical Classifications Software Refined (CCSR)[12] classification system:

- RSP002 - Pneumonia (except that caused by tuberculosis)
- RSP003 - Influenza
- RSP005 - Acute bronchitis
- RSP006 - Other specified upper respiratory infections

Machine learning models were created that use a patient’s historical medical claims data to predict the likelihood they will have an inpatient hospital stay due to one of the above conditions in the next 3 months. The data used was an anonymized 5% sample of the Medicare claims data from 2015 and 2016. This data spanned the transition from International Classification of Diseases version 9 to version 10 (ICD-10) on October 1, 2016. The dataset used to create the model was created by identifying all living members above the age of 18 on 9/30/2016. Only fee-for-service members were included because medical claims histories for other members are not reliably complete. We then excluded all members who had less than 6 months of continuous eligibility prior to 9/30/2016. We also excluded members who lost coverage within 3 months after 9/30/2016, except for those members who lost coverage due to death. Table 1 below summarizes the population selection.

**Table 1:** Features used associated with risk factors identified by CDC and their corresponding CCSR codes

| Cumulative Population Step | Selection Criteria |
| --- | --- |
| 3,114,713 | Total members in the dataset |
| 1,867,879 | Fee for service members with 6 months of continuous coverage prior to 9/30/2016 |
| 1,867,779 | 18 years old or older |
| 1,861,868 | Exclude members with a death date before 9/30/2016 |
| 1,851,519 | Exclude members who lose coverage in the next 3 months not due to death. |

The final dataset is split 80%/20% into train and test sets, with 1,481,654 people in the training set and 369,865 in the test set. The prevalence of the proxy event within the final population was 0.23%.

The labels for the prediction task were created by identifying all patients who had an inpatient visit with an admission date from 10/1/2016 through 12/31/2016 with a primary diagnosis from one of the listed categories. A 3-month delay was imposed on the input features to the model, so that no claims after 6/30/2016 were used to make the predictions. This 3-month delay simulates the delay in claims processing that usually occurs in practical settings and enables the model to be used in realistic scenarios.

### ii. Models

We highlight 3 approaches to building models to help identify individuals who are vulnerable to complications to respiratory infections. All 3 approaches described are machine learning methods created using the same dataset. We have chosen 3 different approaches that represent a tradeoff between accuracy and ease of implementation. For individuals who have access to data, but not the coding background to adopt our model, we hope that the simple model can be easily ported to other systems. For a more robust model, we create a gradient-boosted tree leveraging age, sex, and medical diagnosis history. This model has been made open-source, and can be obtained from github (https://github.com/closedloop-ai/cv19index). Finally, we have created a third model that uses an extensive feature set generated from Medicare claims data along with linked geographical and social determinants of health data. This model is being made freely available through our hosted platform. Information about accessing the platform can be found at https://cv19index.com.

### iii. Logistic Regression

The first approach is aimed at reproducing the high-level recommendations from the CDC website[8] for identifying those individuals who are at risk. They identify risk features as:

- Older adults
- Individuals with heart disease
- Individuals with diabetes
- Individuals with lung disease

To turn this into a model, we extract ICD-10 diagnosis codes from the claims and aggregate them using the CCSR categories. We create indicator features for the presence of any code in the CCSR category. The mapping between the CDC risk factors and CCSR codes is described in Table 2. We start with these features as they give us an ability to quantify the portion of the at-risk population that are encapsulated by the high-level CDC recommendations. In addition to the conditions coming from the recommendations of the CDC, we will look at features that our other modeling efforts surfaced as important and avail those features to the model as well. We also provide gender and age in years, as well as an interaction term between age and the diagnostic features. This simple dataset is used to train a logistic regression model[10]. In addition to the CCSR codes, Table 2 includes the beta coefficients associated with these features in the logistic regression model.

**Table 2:** Features used associated with risk factors identified by CDC and their corresponding CCSR codes

| Feature name | Coefficient | CCSR Code |
| --- | --- | --- |
| Intercept | -5.49 |  |
| Gender male | -0.212 | n/a |
| # of Admissions (12M) | 0.186 | n/a |
| Age | -0.014 | n/a |
| Pneumonia except that caused by tuberculosis | 0.117 | CCSR:RSP002 |
| Other and ill-defined heart disease | -0.111 | CCSR:CIR015 |
| Heart failure | -0.271 | CCSR:CIR019 |
| Acute rheumatic heart disease | -0.008 | CCSR:CIR002 |
| Coronary atherosclerosis and other heart disease | -0.529 | CCSR:CIR011 |
| Pulmonary heart disease | -0.005 | CCSR:CIR014 |
| Chronic rheumatic heart disease | -0.014 | CCSR:CIR001 |
| Diabetes mellitus with complication | -0.273 | CCSR:END003 |
| Diabetes mellitus without complication | -0.496 | CCSR:END002 |
| Chronic obstructive pulmonary disease and bronchiectasis | -0.269 | CCSR:RSP008 |
| Other specified and unspecified lower respiratory disease | -0.020 | CCSR:RSP016 |
| age X Pneumonia | 0.010 | n/a |
| age X Other and ill-defined heart disease | 0.003 | n/a |
| age X Heart failure | 0.009 | n/a |
| age X Acute rheumatic heart disease | 0.003 | n/a |
| age X Coronary atherosclerosis and other heart disease | 0.011 | n/a |
| age X Pulmonary heart disease | -0.000 | n/a |
| age X Chronic rheumatic heart disease | -0.001 | n/a |
| age X Diabetes mellitus with complication | 0.007 | n/a |
| age X Diabetes mellitus without complication | 0.009 | n/a |
| age X Chronic obstructive pulmonary disease and bronchiectasis | 0.013 | n/a |
| age X Other specified and unspecified lower respiratory disease | 0.006 | n/a |

### iv. Gradient Boosted Trees

Our more robust approach uses gradient boosted trees. Gradient boosted trees are a machine learning method that use an ensemble of simple models to create highly accurate predictions[10].

The resulting models demonstrate higher accuracy. A drawback to these models is that they are significantly more complex; however, “by hand” implementations of such models are impractical. Here, we create two variations of the models. The first is a model that leverages information similar to our logistic regression model. A nice feature of gradient boosted trees is that they are fairly robust against learning features that are eccentricities of the training data, but do not extend well to future data. As such, we allow full diagnosis histories to be leveraged within our simpler XGBoost model. In this approach, every category in the full CCSR is converted into an indicator feature, resulting in 559 features. Details about how to connect the full diagnosis history with the open-source model are provided with the open-source version of the model.

We additionally built a model within the ClosedLoop platform. The ClosedLoop platform is a software system designed to enable rapid creation of machine learning models utilizing healthcare data. The full details of the platform are outside the bounds of this paper; however, using the platform allows us to leverage engineered features coming from peer-reviewed studies. Examples are social determinants of health and the Charlson Comorbidity Index[13]. We chose not to include these features within the open-source model, because the purpose of the open-source version is intended to be as accessible as possible for the greater healthcare data science community.

## III. Results and Model Interpretation

We quantify the performance of the CV19 Index using metrics that are standard within the data science community. In particular, we visualize the performance of our model using a receiver operating characteristic graph, see Figure 1. Additionally, the metrics quantifying the effectiveness of our models are presented in Table 3. The performance of both gradient boosted tree models are very similar. The ROC curve demonstrates that as the decision threshold increases, the percentage of the potentially affected population increases at roughly the same rate.The logistic regression model has similar performance at low alert rates. We can see that at a 3% alert rate, the difference in sensitivity is only. 02. The performance at higher alert rates experiences a significant performance disadvantage; however, for most interventions this would be at alert rates higher than is practical.

**Table 3:** Measures of effectiveness for the three models

| Model Type | ROC AUC | Sensitivity at 3% Alert Rate | Sensitivity at 5% Alert Rate |
| --- | --- | --- | --- |
| Logistic Regression | .731 | .214 | .314 |
| XGBoost |  |  |  |
| Diagnosis History + Age | .810 | .234 | .324 |
| XGBoost |  |  |  |
| Full Feature Set | .810 | .231 | .327 |

**Figure 1:**
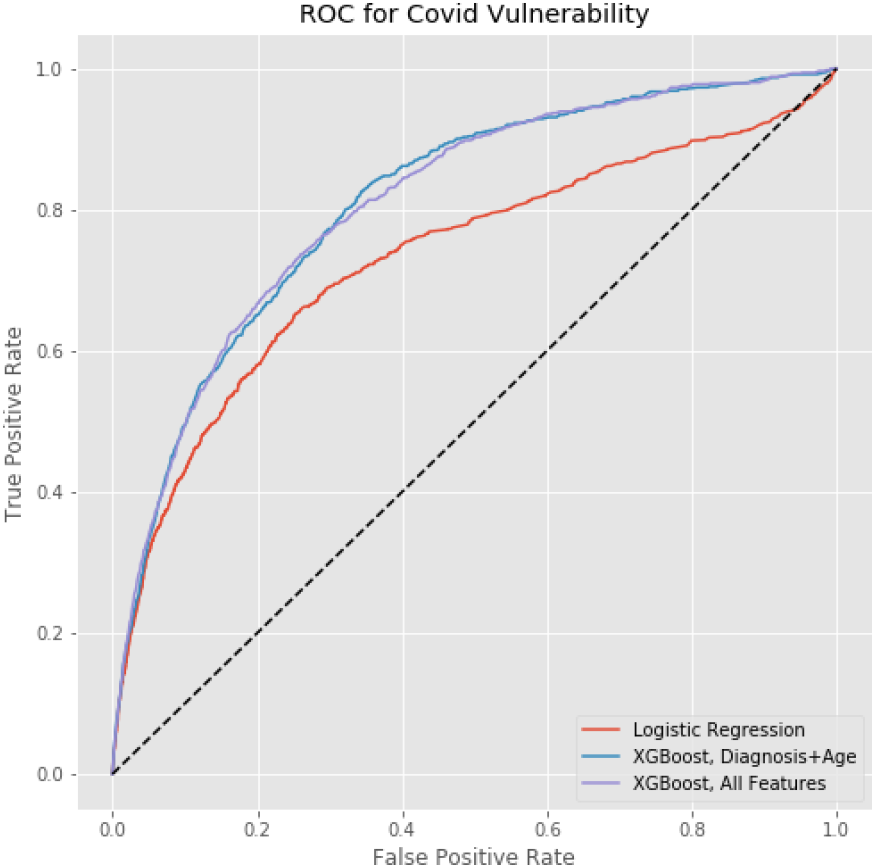
A receiver operating characteristic graph depicting the performance of the 3 models.

## IV. Accessing Models

There are two ways of accessing the models that we are providing. The first is to access the open-source version of our model, which is available at https://github.com/closedloop-ai/cv19index. This model is written in the Python programming language. We have included synthetic data for the purpose of walking individuals through the process of going from tabular diagnosis data to the input format specific for our models. We encourage the healthcare data science community to fork the repository and adapt it to their own purposes. We encourage collaboration from the open-source community, and pull requests will be considered for inclusion in the main branch of the package. For those wishing to use our models within our platform, we are providing access to the COVID-19 model free of charge. Please visit https://closedloop.ai/cv19index for instructions on how to gain access.

## V. Conclusions

This pandemic has already claimed thousands of lives, and sadly, this number is sure to grow. As healthcare resources are constrained by the same scarcity constraints that effect us all, it is important to empower intervention policy with the best information possible. We have provided several implementations of the CV19 Index and means of access for those individuals with varying levels of technical expertise. It is our hope that by providing this tool quickly to the healthcare data science community, widespread adoption will lead to more effective intervention strategies and, ultimately, help to curtail the worst effects of this pandemic.

## Data Availability

The data we used is the CMS Medicare LDS 5% sample for 2015-2016. We cannot distribute this data, but it is available for purchase to to qualified researchers working on projects for the benefit of Medicare beneficiaries.

## VI. Appendix: Full Feature List

We include a full list of features available within our platform. The majority of features are binary variables indicating if a patient has had one type of medical event 15 moths prior to the date of prediction, excluding the 3 most recent months.

**Table 4:**
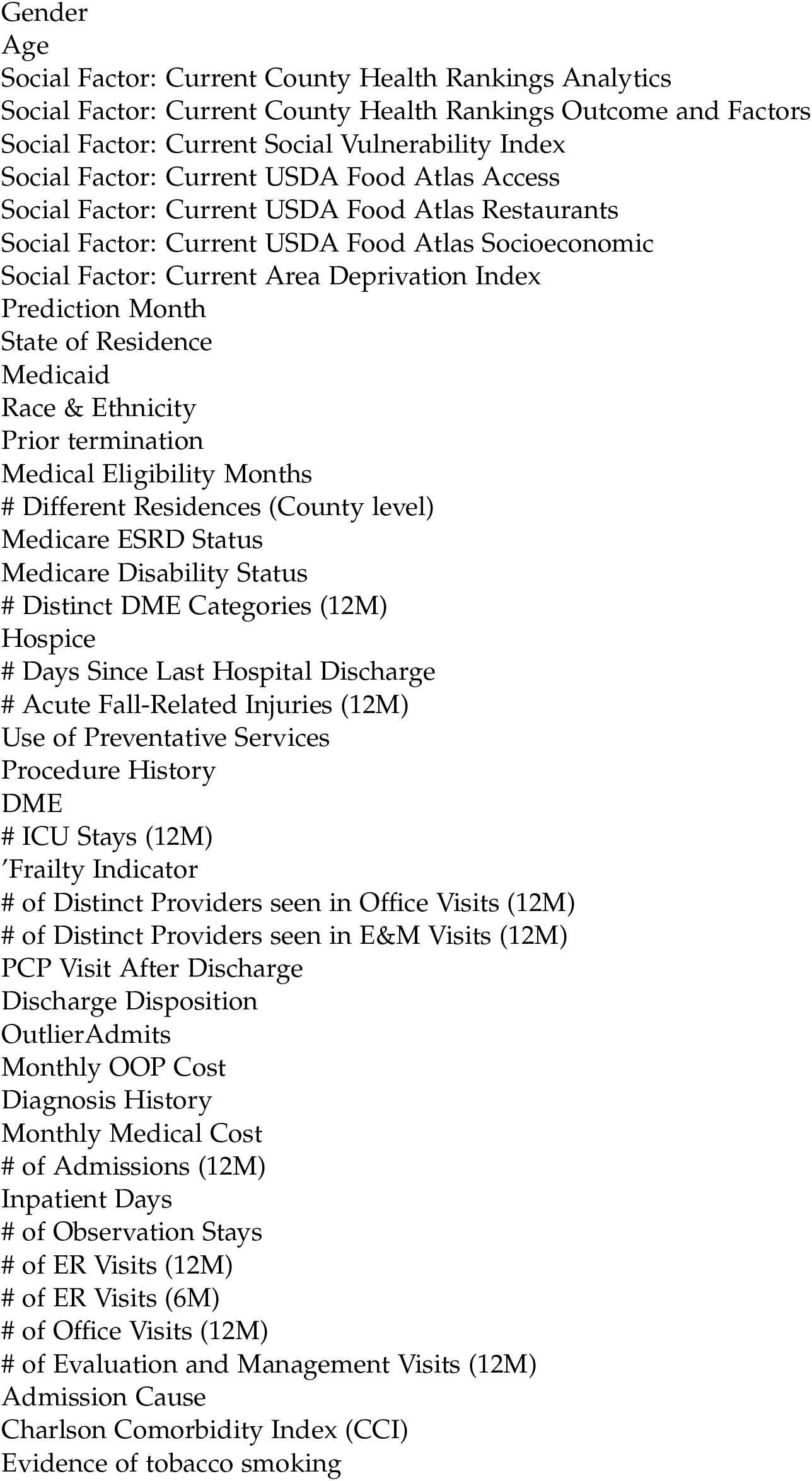
Full feature list available within platform

**Table 5:**
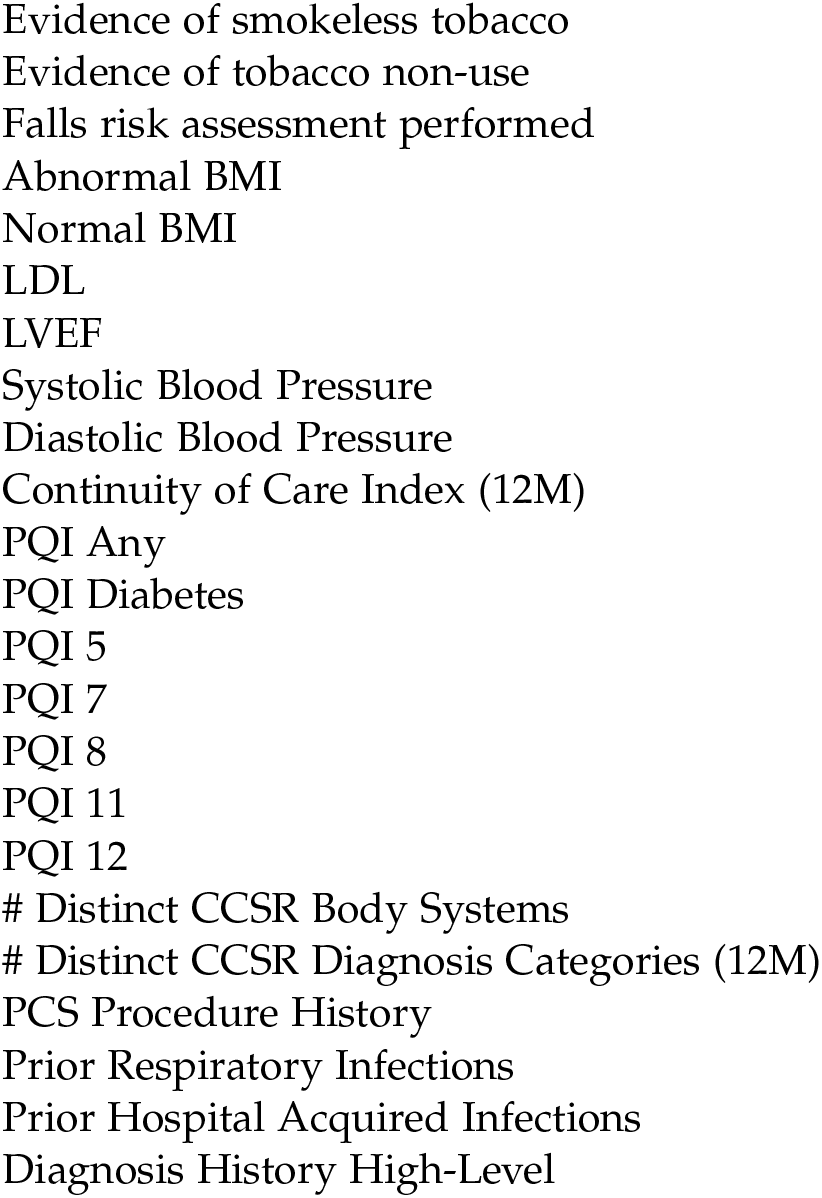
Full feature list available within platform (continued)

## Notes

### Competing Interest Statement

The authors have declared no competing interest.

### Funding Statement

The authors are employed by ClosedLoop.ai, which develops a healthcare data science platform and funded this effort.

